# A Robust, Safe and Scalable Magnetic Nanoparticle Workflow for RNA Extraction of Pathogens from Clinical and Environmental Samples

**DOI:** 10.1101/2020.06.28.20141945

**Authors:** Gerardo Ramos-Mandujano, Rahul Salunke, Sara Mfarrej, Andri Rachmadi, Sharif Hala, Jinna Xu, Fadwa S. Alofi, Asim Khogeer, Anwar M. Hashem, Naif A.M. Almontashiri, Afrah Alsomali, Samir Hamdan, Peiying Hong, Arnab Pain, Mo Li

## Abstract

Diagnosis and surveillance of emerging pathogens such as SARS-CoV-2 depend on nucleic acid isolation from clinical and environmental samples. Under normal circumstances, samples would be processed using commercial proprietary reagents in Biosafety 2 (BSL-2) or higher facilities. A pandemic at the scale of COVID-19 has caused a global shortage of proprietary reagents and BSL-2 laboratories to safely perform testing. Therefore, alternative solutions are urgently needed to address these challenges. We developed an open-source method called <u>M</u>agnetic-nanoparticle-<u>A</u>ided <u>V</u>iral <u>R</u>NA <u>I</u>solation of <u>C</u>ontagious <u>S</u>amples (MAVRICS) that is built upon reagents that are either readily available or can be synthesized in any molecular biology laboratory with basic equipment. Unlike conventional methods, MAVRICS works directly in samples inactivated in acid guanidinium thiocyanate-phenol-chloroform (e.g., TRIzol), thus allowing infectious samples to be handled safely without biocontainment facilities. Using 36 COVID-19 patient samples, 2 wastewater samples and 1 human pathogens control sample, we showed that MAVRICS rivals commercial kits in validated diagnostic tests of SARS-CoV-2, influenza viruses, and respiratory syncytial virus. MAVRICS is scalable and thus could become an enabling technology for widespread community testing and wastewater monitoring in the current and future pandemics.

## Introduction

Testing for COVID-19 is vital for monitoring and mitigating the spread of SARS-CoV-2 and for safely restarting the normal economy. To date, molecular diagnosis of COVID-19 predominantly relies on detection of SARS-CoV-2 RNA using real-time reverse transcription polymerase chain reaction (rRT-PCR) assays, such as those approved by the US Centers for Disease Control and Prevention (US CDC)(1). As SARS-CoV-2 spreads globally, it also accumulates approximately 1 to 2 single nucleotide variants (SNVs) in the 29,903 bp genome per month(2). The emergence of new strains could have serious implications in the efficacy of diagnostic tests and success of vaccines. For example, 87 of 2816 genomes sampled between Jan and May 2020 have the T28688C SNV (GISAID, https://nextstrain.org/) that alters the sequence of the binding site of the forward primer of the CDC N3 rRT-PCR assay(1), potentially compromising its effectiveness. Thus, continued surveillance of the evolution and geographic distribution of viral strains by high-throughput sequencing (3,4) is another pillar of public health measures to combat COVID-19.

Both rRT-PCR testing and high-throughput sequencing of SARS-CoV-2 require RNA extraction from nasopharyngeal swab samples. In the clinic, swabs are collected in viral transport media (VTM) and, if necessary, transported following specific cold-chain biological substances transport guidelines(1) for RNA extraction. The US CDC recommends several commercially available RNA extraction kits(1). Fully automated diagnostic systems (e.g., Roche cobas® 6800 and 8800) that perform all steps from RNA extraction to rRT-PCR without human intervention are also popular among diagnostic laboratories. Commercial kits and procedures typically yield consistent quality RNA and are easy to use, but come with a high price tag. Moreover, the availability of commercial proprietary reagents is seriously affected by the disruption of the global supply chain caused by the COVID-19 pandemic. The high cost and low availability of proprietary reagents impose a bottleneck on testing capacities in rich and poor countries alike. Additionally, monitoring pathogens in wastewater is an important public health measure, and it requires methods that satisfy the biosafety requirements of handling unknown infectious agents and can overcome the low virus concentration and PCR inhibitors that are ubiquitous in wastewater. Therefore, there is great incentive to develop alternative methods that only require locally available and inexpensive chemicals, are simple to perform, and rival the performance of commercial kits. Besides alleviating supply shortage, the alternative methods should ideally eliminate the risk of handling live viruses, thus lowering the strict biosafety and biosecurity requirements(5) on testing facilities. Any self-build RNA extraction method that satisfies the above-mentioned criteria can help increase testing capacity not only in clinical laboratories but also in rural healthcare facilities, university laboratories and field testing sites.

RNA isolation by acid guanidinium thiocyanate-phenol-chloroform extraction (AGPC)(6) (sold as TRIzol by Invitrogen or TRI Reagent by Sigma-Aldrich) has been successfully used in life sciences laboratories around the world for nearly four decades. It requires widely available chemicals at a low cost. The AGPC methods has been found to match the performance of commercial kits and automated systems in SARS-CoV-2 rRT-PCR detection(7,8). In these studies, swabs were first collected in VTM or cell culture media, which were then used in AGPC RNA isolation. This workflow necessitates handling of live viruses and requires Biosafety Level 2 (BSL2) facilities.

We hypothesized that it should be possible to collect swabs directly in AGPC, which achieve two goals: 1) completely inactivation of any infectious agent by AGPC so that the downstream procedures (e.g., transportation, RNA isolation, rRT-PCR, and sequencing) can be carried without BSL2 requirements, and 2) preservation of RNA integrity by denaturing nucleases.

However, the AGPC method as is commonly practiced has several drawbacks that make it unsuitable for high-volume testing. It requires extensive manual pipetting of hazardous chemicals and multiple centrifugation steps, which increase the risk of human errors and personnel injury especially when the sample number is large. Solid-phase reversible immobilization (SPRI) of nucleic acid on magnetic nanoparticles (MNPs) offers a simple and elegant alternative to centrifuge- or column-based methods(9). Nucleic acid (e.g. RNA) reversibly binds to functionalized MNPs under dehydrating conditions and can be separated from contaminants in solution by a strong magnet. This allows fast and thorough washes to eliminate inhibitors of downstream molecular biology reactions and yields high quality RNA for PCR and high-throughput sequencing. Because it requires no centrifugation and only low-cost materials, the MNP-based RNA extraction is inherently scalable and amenable to automation. Although the combination of the AGPC and SPRI technologies would be obviously advantageous in consideration of reagent availability, cost, biosafety and ease-of-use, development of AGPC compatible MNP-based RNA extraction protocols has been limited.

Here we developed the <u>M</u>agnetic-nanoparticle-<u>A</u>ided <u>V</u>iral <u>R</u>NA <u>I</u>solation of <u>C</u>ontagious <u>S</u>amples (MAVRICS) workflow (**Fig. 1A**). MAVRICS only requires widely available and low-cost materials and can be self-assembled in a basic laboratory setting. It is compatible with AGPC inactivated samples to alleviate the shortage of commercial kits, lower biosafety risks, and enable sample and scalable sample preparation. MAVRICS performed on par or better than commercial RNA extraction kits in rRT-PCR detection of SARS-CoV-2, influenza viruses and respiratory syncytial virus in various clinical and environmental samples.

**Figure 1.**
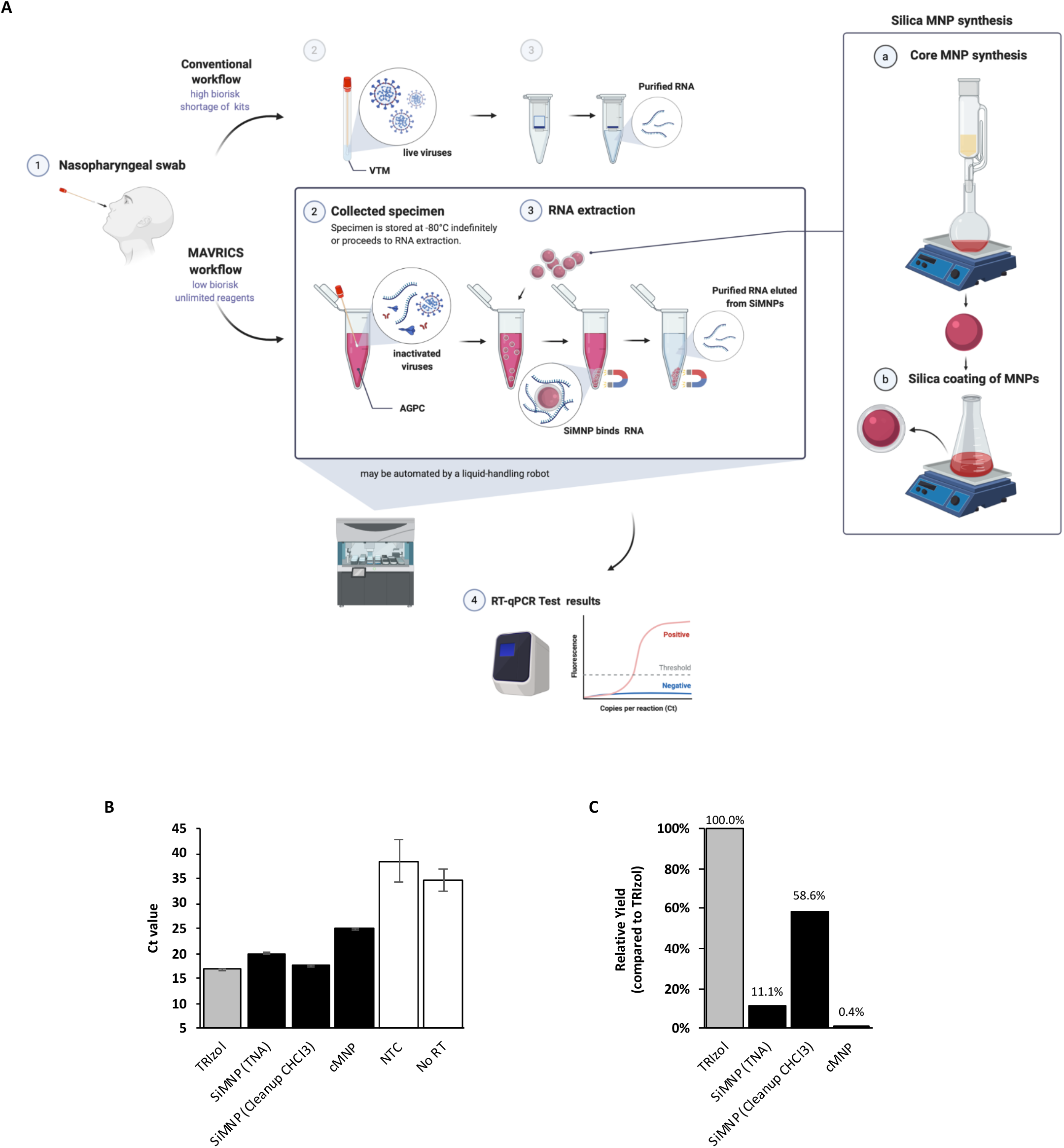
Silica-coated magnetic nanoparticles can isolate RNA directly from AGPC inactivated samples. **A**. A schematic comparison of the conventional and magnetic nanoparticle-aided viral RNA isolation of contagious samples (MAVRICS) workflow for COVID-19 nasopharyngeal swabs. **B-C**. Silica magnetic nanoparticles (SiMNP) synthesized using an open-source protocol are able to isolate viral RNA. RNA was extracted from contrived SARS-CoV-2 saliva samples inactivated in TRIzol using different methods. The TRIzol Reagent protocol was used as a control. After RNA extraction rRT-PCR was conducted with the US CDC 2019-nCoV_N3 assay. **B**. Ct values, and **C**. Viral RNA yield relative to TRIzol extraction. AGPC: acid guanidinium thiocyanate-phenol-chloroform. TNA: total nucleic acid extraction protocol, cMNP: carboxylated MNPs, NTC: no template control, No RT: no reverse transcriptase control. N=3, error bars represent standard errors.

## Material &Methods

### Clinical samples

Contrived SARS-CoV-2 saliva samples were prepared by mixing 1000 μl of TRIzol, 100 μl of saliva from a health volunteer, and 5 μl of in vitro transcribed SARS-CoV-2 N gene RNA (nt28,287-29,230 in NC_045512.2, 10^8^ copies/ul). Anonymized RNA samples were obtained from Ministry of Health (MOH) hospitals in the western region in Saudi Arabia. The use of clinical samples in this study is approved by the institutional review board (IRB# H-02-K-076-0320-279) of MOH and KAUST Institutional Biosafety and Bioethics Committee (IBEC). Oropharyngeal and nasopharyngeal swabs were carried out by physicians and samples were steeped in 1 mL of TRIzol (Invitrogen Cat. No 15596018) to inactivate virus during transportation. The Respiratory (21 targets) control panel (Microbiologics Cat. No 8217) was used as controls in rRT-PCR assays.

### Wastewater samples and virus concentration

One liter of raw sewage is individually sampled at 9 AM and 4 PM on 7 June 2020 from the equalization tank of wastewater treatment plant operated within KAUST. The sewage from both time-points are then mixed together to constitute a composite sample. A 300-500 ml of raw sewage was concentrated by using an electronegative membrane in the present of cation which has been described previously (Reference 14). Briefly, 2.5 M MgCl_2_ was added to the water samples to obtain a final concentration of 25 mM. The samples were subsequently passed through the electronegative filter (cat. no. HAWP-090-00; Merck Millipore, Billerica, MA) attached to a glass filter holder (Merck Millipore, Cat no. XX1009020). Magnesium ions were removed by passing 200 mL of 0.5 mM H2SO4 (pH 3.0) through the filter, and the viruses were eluted with 10 mL of 1.0 mM NaOH (pH 10.8). The eluate was recovered in a tube containing 50 μL of 100 mM H2SO4 (pH 1.0) and 100 μL of 100× Tris–EDTA buffer (pH 8.0) for neutralization. The samples were further concentrated using a Centripep YM-50 (Merck Millipore) to obtain a final volume of 600–700 μL.

### Magnetic nanoparticle synthesis, silica coating and RNA extraction protocol

Core magnetic nanoparticle synthesis and silica coating of MNPs were done following published protocols (Protocols 1.1 and 2.1 in reference 12). Bis-Tris and Tris binding buffer (Bis-Tris buffer and Tris buffer): To prepare 50 ml of binding buffer dissolve 14.33 g of Gu-HCl (3 M final concentration) and 104.6 mg of Bis-Tris or 60.5 mg of Tris Base (10 mM final concentration) in 45 ml of Ethanol 100%. Adjust pH with HCl (pH < 6.5) and adjust the volume with water to 50 ml. Bis-Tris or Tris binding buffer (200 µl) and SiMNP (40 μl) were added to samples (200 µl), and shake for 5 min at 1300 rpm. The tubes were settled on a magnetic stand and the cleared supernatant was removed. Then, TRIzol reagent (200 µl) and binding buffer (200 µl) were added and the tubes were vortexed and settled on a magnetic stand and the cleared supernatant was removed. 90% Ethanol (400 µl) was added and the tubes were vortexed and settled on a magnetic stand. The supernatant was then removed. Ethanol washing was repeat three more times for a total of four washes. The beads were dried on a heat block at 50 °C for ∼20 min. To elute the RNA 40 μl of nuclease-free water was added and mixed at 1300 rpm at RT for 5 min.

Finally, the tube was settled on a magnetic stand and the eluted RNA transferred to a new tube. A detailed supplementary protocol 1 can be found in online supplementary materials. A step-by-step protocol is also available at url: https://dx.doi.org/10.17504/protocols.io.bik4kcyw.

### RNA extraction by commercial methods

Total RNA extraction of the samples was performed following instructions as described in the CDC EUA-approved protocol using the Direct-Zol RNA Miniprep kit (Zymo Research Cat. No R2070) or TRIzol reagent (Invitrogen Cat. No 15596026) following the manufacturers’ instructions. RNA extraction with RNAClean XP beads was done following manufacturer instruction. Viral RNA were extracted from the concentrated raw sewage by using QIAamp viral RNA mini kit (Qiagen, cat no: 52906) following manufacture instruction. A 140 μL of concentrated raw sewage was used to obtain a final elution of 80 μL. The RNA was stored in – 20 °C freezer until further use.

### Reverse transcription

Reverse transcription of RNA samples was done using either NEB ProtoScript II reverse transcriptase (NEB Cat. No M0368) or Invitrogen SuperScript IV reverse transcriptase (Thermo Fisher Scientific Cat. No 18090010), following protocols provided by the manufacturers. After reverse transcription, 5 units of thermostable RNase H (New England Biolabs Cat. No M0523S) was added to the reaction, which was incubated at 37 °C for 20 min to remove RNA. The final reaction was diluted 10-fold to be used as templates in RPA. All of the web-lab experiments in this study were conducted in a horizontal flow clean bench to prevent contaminations. The bench was decontaminated with 70% ethanol, DNA*Zap* (Invitrogen, Cat no. AM9890) and RNase *AWAY* (Invitrogen, Cat no. 10328011) before and after use. The filtered pipette tips (Eppendorf epT.I.P.S.® LoRetention series) and centrifuge tubes (Eppendorf DNA LoBind Tubes, Cat. No 0030108051) used in this study were PCR-clean grade. All of the operations were performed carefully following standard laboratory operating procedures.

### Real-time PCR

Real-time PCR assays for SARS-CoV-2 were purchased from IDT (Cat. No 10006770). Real-time PCR analysis of SARS-CoV-2 sequences were analyzed on a CFX384 Touch Real-Time PCR Detection System (Biorad) using the following program: 50°C for 2 min, 95°C for 2 min followed by 45 cycles of 95°C for 5 sec and 59°C for 30 sec. Real-time PCR assays for Influenza A, B/RSV were purchase from IDT (Cat. No 1079729) and used per the manufacturer’s recommendation. For Influenza and RSV assays, the following program was used: 50°C for 2 min, 55°C for 120 sec, 60°C for 360 sec, 65°C for 240 sec, followed by 5 cycles 95°C for 5 sec and 55°C for 30 sec, and then 45 cycles of 91°C for 5 sec and 58°C for 25 sec. MNV and PMMoV real-time PCR assay was conducted using the primer and probes which described previously(10,11).

## Results

### Silica-but not carboxyl-coated magnetic nanoparticles can isolate RNA directly from AGPC inactivated samples

MNPs can be functionalized with either a carboxyl or silica coating to bind nucleic acids(12). Carboxylated MNPs (cMNPs) are available commercially (e.g., RNAClean XP from Beckman Coulter) and widely used in molecular biology workflows such as PCR cleanup and sequencing library preparation. Unfortunately, cMNPs (in the form of RNAClean XP) failed to recover detectable RNA from AGPC solutions (in the form of TRIzol) spiked with high quality total RNA from human cells, while the conventional AGPC method based on organic phase separation and centrifugation recovered ∼45% of input RNA. On the other hand, cMNPs were capable of 84-96% recovery when the same RNA was spiked in water, suggesting that AGPC interferes with RNA binding onto cMNPs. Silica magnetic nanoparticles (SiMNP) have been used to extract total nucleic acid from samples lysed and inactivated in AGPC without centrifugation and phase separation(12). Since commercial SiMNPs are expensive and difficult to procure during the COVID-19 crisis, we synthesized SiMNP from scratch using a published open-source protocol(12). The synthesis took ∼14 hours with 3 hours hands-on time and required only base chemicals, a strong magnet, and standard lab equipment (**Fig. 1A, Supplementary Fig. 1A-D**). In our case, all materials were locally available (**Table 1**). One synthesis yielded enough SiMNPs for 5,000-10,000 extractions, and the material cost was miniscule. Another benefit of SiMNP is its chemical inertness. Our SiMNPs have been stored at room temperature for 6 weeks at the time of writing without noticeable change in performance.

**Table 1.** List of Materials for SiMNP Synthesis, RNA Extraction and rRT-PCR. The vendors and catalog numbers are those used in this study. Alternative sources can also be used.

| REAGENT | SUPPLIER | CATALOG NUMBER |
| --- | --- | --- |
| Iron (II) chloride tetrahydrate ≥98% (FeCl<sub>2</sub> · 4 H<sub>2</sub>O) | VWR Chemicals | 13478-10-9 |
| Iron (III) chloride, anhydrous, extra pure (FeCl<sub>3</sub>) | Fisher Scientific | 10224390 |
| Sodium hydroxide, ≥99% (NaOH) | Sigma Aldrich | 306576-500G |
| Hydrochloric Acid (36.5 to 38.0%) | Fisher Scientific | A144-500 |
| Ammonia solution (NH<sub>4</sub>OH, 25%) | Fisher Scientific | 10642251 |
| Ethanol absolute ≥99.8% | VWR Chemicals | 20821.330 |
| Tetraethyl orthosilicate (≥99%) (GC) | Sigma Aldrich | 78-10-4 |
| 2,2-Bis(hydroxymethyl)-2,2',2''-nitrilotriethanol /Bis-Tris (C<sub>8</sub>H<sub>19</sub>NO<sub>5</sub>) | Gold Biotechnology | B-020-500 |
| Guanidinium chloride / Gu-HCl (CH<sub>5</sub>N<sub>3</sub> · HCl) | Fisher Scientific | BP178-1 |
| Tris Base | Promega Corporation | H5135 |
| Tween 20 | Sigma Aldrich | P7949 |
| TRIzol reagent | Life Technologies | 15596018 |
| SuperScript™ IV Reverse Transcriptase | Invitrogen | 18090010 |
| RNase OUT | Invitrogen | 10777-019 |
| TaqMan™ Fast Advanced Master Mix | Invitrogen | 4444556 |
| RNase H | New England BioLabs | M0297L |
| 2019-nCoV Kit | Integrated Device Technology (IDT) | 10006605 |
| Influenza/RSV qPCR Assay | Integrated Device Technology (IDT) | 1079729 |
| Direct-Zol RNA Miniprep kit | Zymo Research | R2070 |
| QIAamp viral RNA mini kit | Qiagen | 52906 |
| ProtoScript II reverse transcriptase | New England BioLabs | M0368 |

We first tested if SiMNPs could isolate RNA from contrived SARS-CoV-2 saliva samples (see methods) inactivated in AGPC (in the form of TRIzol). As previously reported, SiMNPs were able to isolate RNA directly from TRIzol using the total nucleic acid extraction protocol (hereafter referred to as TNA protocol) described in(12). We used the US CDC 2019-nCoV_N3 rRT-PCR assay to quantitate the recovery of SARS-CoV-2 sequences. SiMNPs coupled with the TNA protocol resulted an increase of 3.1 in Ct value compared to the official TRIzol Reagent protocol, which means a 11.1% yield of viral RNA relative to the AGPC method (**Fig. 1B-C**). In contrast, RNA received by the cMNP (RNAClean XP) methods was negligible (**Fig. 1B-C**). Interestingly, the yield of SiMNPs improved when the sample in TRIzol was first phase separated by chloroform and the aqueous phase was used in combination with an enzymatic reaction cleanup protocol described in(12)(cleanup CHCl_3_ protocol, **Fig. 1B-C**). However, this modification defeated the purpose of using SiMNPs to simplify the workflow. Together, these results showed that SiMNPs could isolate viral RNA directly from AGPC inactivated samples, but existing SiMNP protocols significantly underperformed compared to the AGPC method, thus reducing the sensitivity of diagnostic tests.

### Development of a SiMNP-based protocol to maximize viral RNA recovery

Next, we aimed to develop an efficient SiMNP-based RNA extraction protocol using the contrived SARS-CoV-2 samples and US CDC 2019-nCoV_N1 and N3 rRT-PCR assays. Increasing the amount of SiMNPs 2.5 times significantly improved the recovery of both the TNA and cleanup CHCl_3_ protocols. We also noticed an improvement by washing the SiMNPs once with TRIzol and RNA binding buffer (1:1), presumably further removing RNases. Nonetheless, none of these protocols could better the TRIzol Reagent protocol (**Fig. 2A-B, Supplementary Fig. 2B-C**). Since the cleanup CHCl_3_ protocol had consistently outperformed the TNA protocol, we suspected that the RNA binding buffer(12) in the TNA protocol might not be optimal. Indeed, after adding buffering agents (Tris-HCl or Bis-Tris, pH6.5) to the RNA binding buffer and increasing its guanidinium chloride concentration to 3M, the yield of RNA doubled (**Fig. 2A-B, Supplementary Fig. 2B-C**).

**Figure 2.**
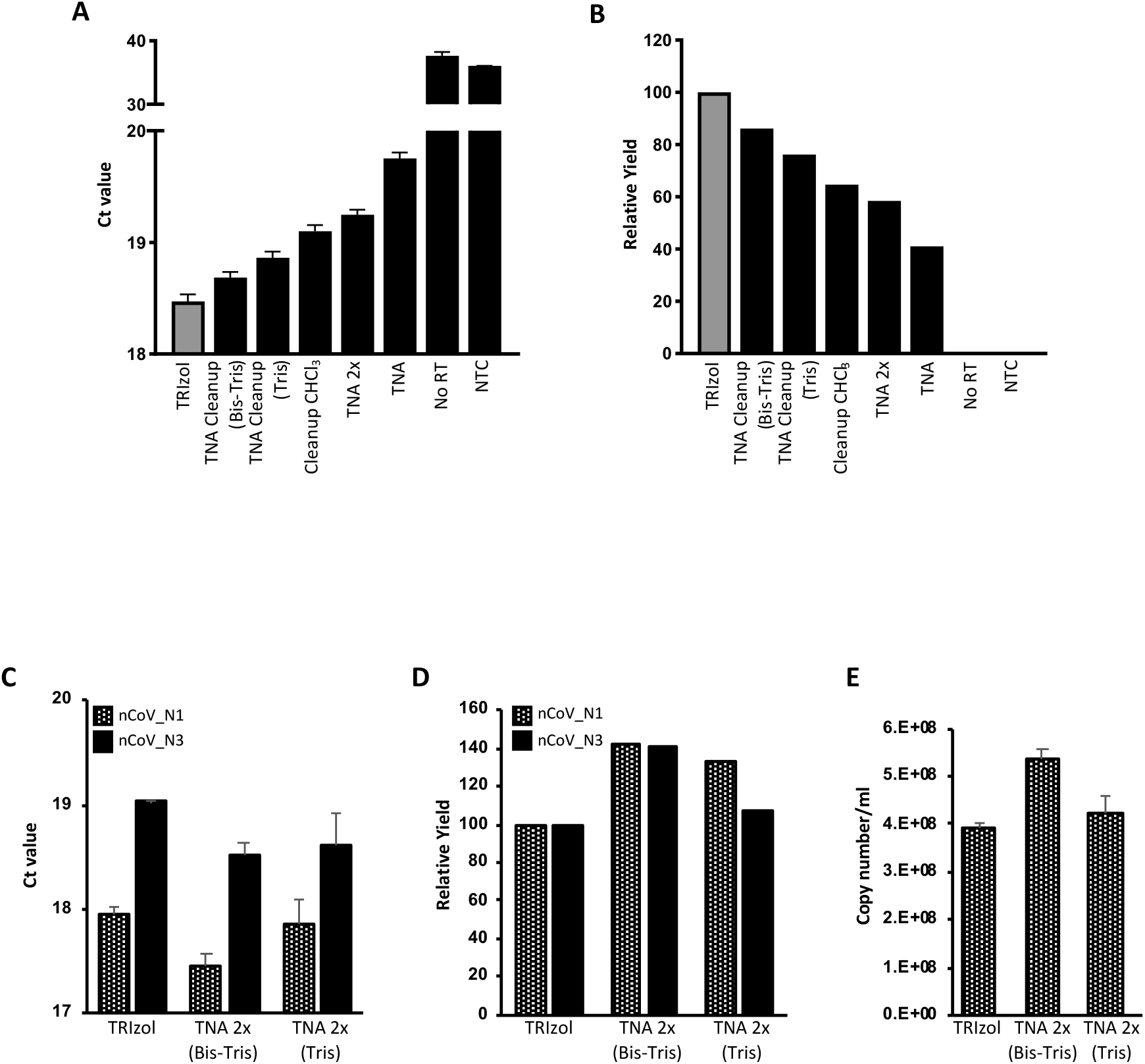
Optimization of SiMNP protocol to maximize viral RNA recovery. **A and B**. The SARS-CoV-2 RNA recovery of various SiMNP protocols was compared using the 2019-nCoV_N3 rRT-PCR assay. **A:** Ct values. **B:** viral RNA yield relative to TRIzol extraction. **C and D**. The SARS-CoV-2 RNA recovery of the optimized SiMNP protocols was analyzed using the 2019-nCoV_N1 and N3 rRT-PCR assays. **C:** Ct values. **D:** viral RNA yield relative to TRIzol extraction. **E**. Copy number of SARS-Cov-2 RNA in the original sample calculated by the standard curve method. Tris: Tris-HCl pH6.5 buffer. Bis-Tris: Bis-Tris, pH6.5 buffer, TNA 2x: TNA protocol with an additional TRIzol wash. N=3, error bars represent standard errors.

We combined the modifications, i.e., the additional wash step and new binding buffers, that improved the recovery of viral RNA by SiMNPs and showed that they outperformed the TRIzol Reagent protocol as judged by both the N1 and N3 rRT-PCR assays (TNA 2X Bis-Tris or Tris, **Fig. 2C-D**). The number of SARS-CoV-2 RNA molecules captured by the SiMNP-TNA 2X Bis-Tris or SiMNP-TNA 2X Tris protocol was estimated by the standard curve method to be very close to the input value (**Fig. 2E**). Similar results were obtained using an independent synthesis of SiMNPs, proving the robustness of the protocols (**Supplementary Fig. 2E-G**). Because of the high recovery of SARS-CoV-2 viral RNA, we name the method (SiMNP coupled to the TNA 2X Bis-Tris protocol) Magnetic-nanoparticle-Aided Viral RNA Isolation of Contagious Samples (MAVRICS).

### Comparing performance of MAVRICS and commercial RNA extraction kits in SARS-CoV-2 rRT-PCR diagnostic panel using clinical samples

We next compared MAVRICS with commercial kits using real-world COVID-19 swab samples obtained in hospitals in the Western Region of Saudi Arabia. These swabs were directly inactivated in TRIzol at the time of collection. Equal aliquots of twelve COVID-19 samples lysed in TRIzol (S659-S670) were extracted using the MAVRICS and TRIzol Reagent protocol respectively. The Ct values obtained by both methods were highly concordant (correlation coefficient=0.96, **Fig. 3A**). MAVRICS on average provided a reduction in Ct value (0.54±0.41, **Fig. 3B**). We further used these and additional 24 samples to compare MAVRICS with the DIRECT-zol RNA kit, which is a proprietary column-based method for RNA purification from TRIzol or similar AGPC reagents. The correlation coefficient between the Ct value of MAVRICS and DIRECT-zol was 0.22 and 0.13, for the US CDC 2019-nCoV_N1 and N2 rRT-PCR assays, respectively (**Fig. 3C, Supplementary Fig. 3A)**. Again, MAVRICS on average provided a reduction in Ct value for both N1 and N2 assays (N1: −0.98±0.92, N2: −0.31±1.0, **Fig. 3D, Supplementary Fig. 3B**). The virus load in the 36 samples were estimated to range between 6.84×10^3^ and 7.52×10^7^ copies/ml (**Supplementary Fig. 3C**).

**Figure 3.**
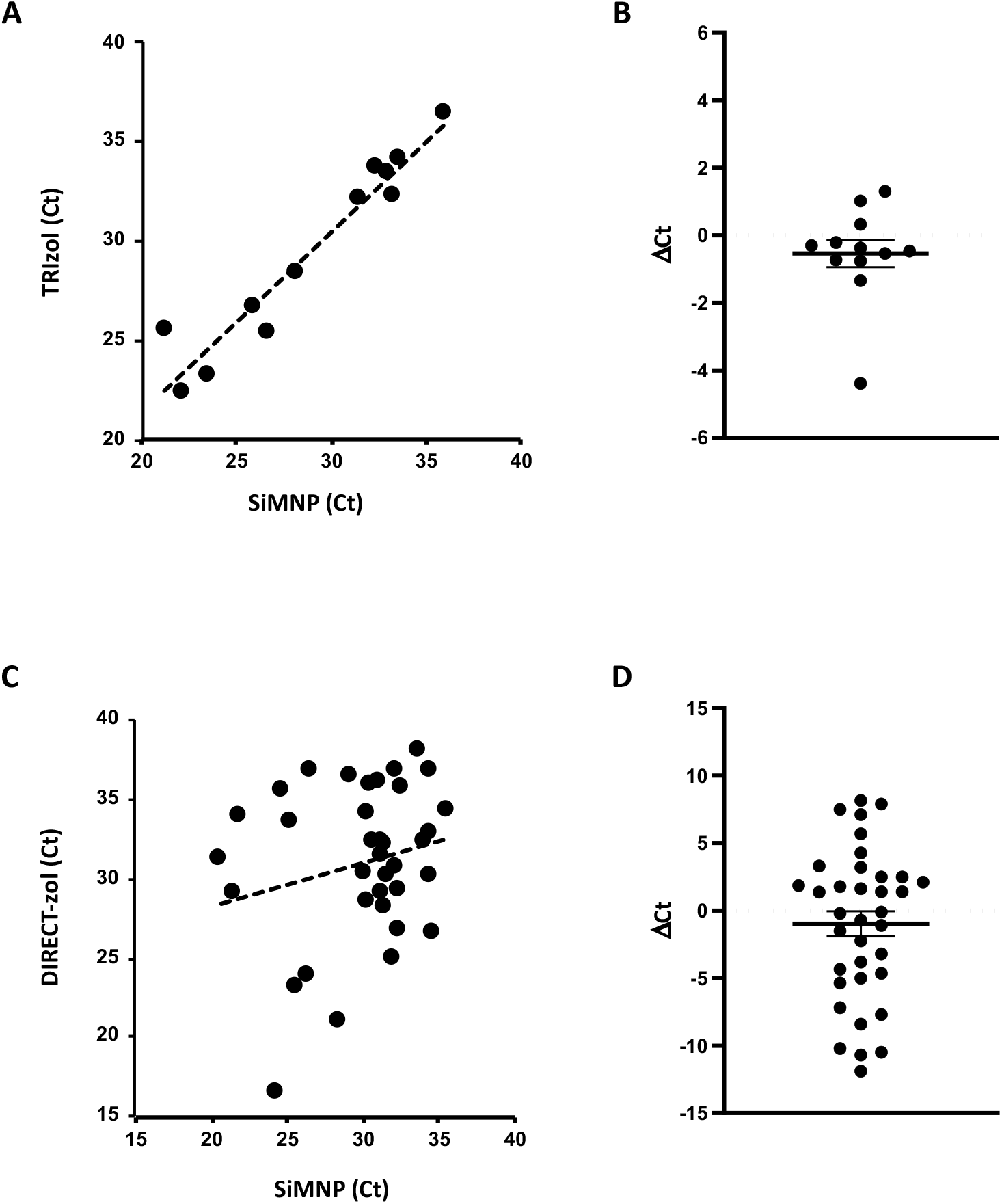
Comparison between MAVRICS and commercial kits in RNA extraction from CIVID-19 clinical samples. RNA extraction was done from 12 samples using MAVRICS and the TRIzol Reagent protocol (**A and B**, 2019-nCoV_N3 assay) or 36 samples using MAVRICS and the DIRECT-zol protocol (**C and D**, 2019-nCoV_N3 assay). The graphs show the correlation between Ct values (**A and C**) and ΔCt values (**B and D**, mean and standard errors are shown).

### MAVRICS is compatible with detection of SARS-CoV-2 and other viruses in wastewater samples

Since the first reports of SARS-CoV-2 shedding in stool (13,14), the presence of the virus has been confirmed in municipal wastewater, sometimes even before the first confirmed cases in the community (15). This suggests that wastewater surveillance could be effective for monitoring the total COVID-19 case load (including asymptomatic cases) in the population.

Detecting pathogens by rRT-PCR in wastewater requires methods that satisfy the biosafety requirements of handling unknown infectious agents and can overcome the low virus concentration and PCR inhibitors that are ubiquitous in wastewater. MAVRICS could be a safe and easy-to-implement workflow to extract viral RNA in wastewater. We first tested the recovery of known quantities of SARS-CoV-2 RNA and intact murine noroviruses (MNVs) spiked in wastewater concentrate, in which viral particles in 250 ml raw sewage were concentrated on electronegative membranes followed by ultrafiltration with Centripep YM-50 to a final volume of 700 ul (16). The wastewater concentrate was first inactivated by 10X volume of TRIzol and extracted using MAVRICS. The result showed an 88% recovery of the input SARS-CoV-2 RNA (**Fig. 4A**). The amount of norovirus RNA captured by the SiMNPs was almost identical to that by the conventional Qiagen RNA purification kit (**Fig. 4B**).

**Figure 4.**
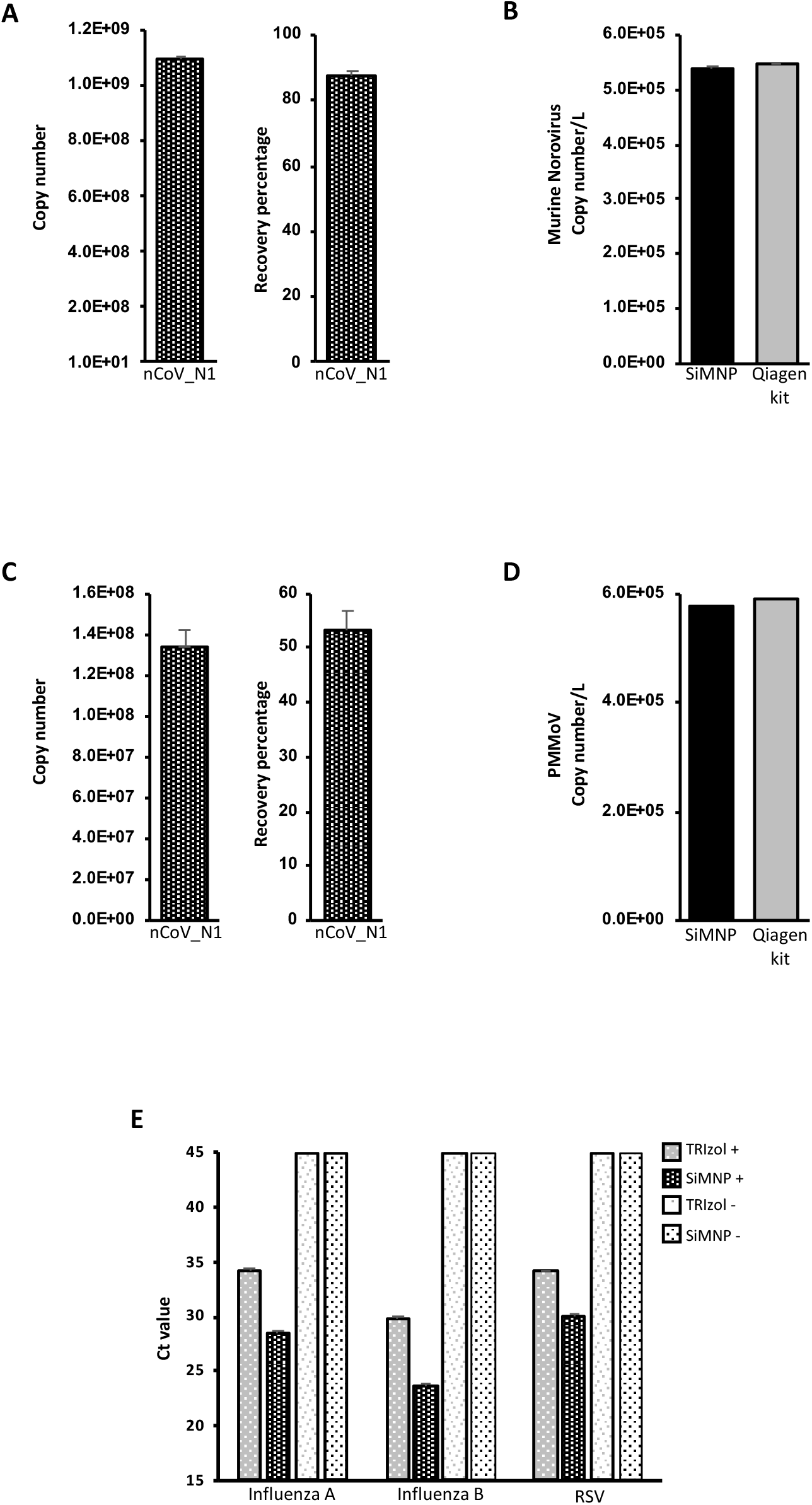
MAVRICS is compatible with wastewater surveillance and detection of other human pathogenic viruses. RNA was extracted by MAVRICS or by the QIAamp viral RNA mini kit (Qiagen kit) from **A-B:** wastewater concentrate samples spiked with SARS-CoV-2 RNA and intact murine noroviruses (MNVs). **C-D:** wastewater biomass immobilized on electronegative membranes with SARS-CoV-2 RNA spike-in. In **A** and **C** the SARS-CoV-2 RNA copy numbers were calculated by the standard curve method. The recovery of viral RNA was compared to the input amount. In **B** and **D** MNV and PPMoV copy numbers in the original sample were compared between MAVRICS and Qiagen kit. **E**. RNA was extracted using MAVRICS or the TRIzol Reagent protocol from a human respiratory pathogens control panel (influenza A and B viruses, and respiratory syncytial virus (RSV)). The Ct was obtained using a clinical diagnostic rRT-PCR panel to quantitate the viruses. TRIzol+ and SiMNP+: positive control panel containing pathogens. TRIzol- and SiMNP-: blank control panel without microorganism. N=3 (**A** and **D**), 6 (**B** SiMNP), 2 (**B** Qiagen kit), 6 (**C**), and 5 (**E**). Error bars represent standard errors.

We further simplified the preparation of wastewater by using TRIzol to inactivate and lyse the sewage biomass (including viral particles) immobilized on the electronegative membranes, followed by RNA extraction by MAVRICS. Again, the spike-in SARS-CoV-2 was efficiently recovered (**Fig. 4C**), and the amount of pepper mild mottle virus (PPMoV, ubiquitous in wastewater) captured by the SiMNPs was almost identical to that by the conventional QIAamp viral RNA mini kit (**Fig. 4D**).

### MAVRICS is compatible with detection of influenza A/B and respiratory syncytial virus

Lastly, we validated the MAVRICS method for detection of other human pathogenic viruses than SARS-CoV-2. A commercial human respiratory pathogens control panel that contains influenza A and B viruses, and respiratory syncytial virus (RSV) was lysed in TRIzol and used for RNA extraction by MAVRICS. We then used a clinical diagnostic rRT-PCR panel to quantitate the viruses. Interestingly, influenza A, influenza B and RSV were readily detectable in samples extracted using SiMNPs, but the Ct value of the same pathogens lagged by 4.08, 6.03 and 5.57, respectively, for samples extracted using the TRIzol Reagent protocol (**Fig. 4E**). No virus was detected in blank controls extracted either by SiMNPs or TRIzol (**Fig. 4E**).

## Discussion

We described an SiMNP-based RNA extraction workflow, MAVRICS, that is compatible with pathogen detection in clinical and environmental samples. All reagents used in MAVRICS are either readily available or can be synthesized in any molecular biology laboratory with basic equipment. The longest preparation step is the synthesis and silica coating of MNPs, which can be done overnight with ∼ 3-hour hands-on time. The material cost for one synthesis is inconsequential yet can support thousands of RNA extractions. Because MAVRICS works for samples inactivated and preserved in AGPC (e.g., TRIzol), it allows potentially infectious samples to be handled safely without special biocontainment facilities. Importantly, MAVRICS matches, and exceeds in many cases, the performance of commercial proprietary reagents using established molecular diagnostic tests of SARS-CoV-2, influenza viruses, and RSV. These tests entail molecular biology reactions that require high quality input RNA. This suggests that the RNA produced by MAVRICS is free of contaminants and maintains good integrity. It will be of interest to study if MAVRICS is compatible with other molecular biology techniques, such as next-generation sequencing (NGS), in the future. Since NGS library preparation uses similar reactions, including reverse transcription and PCR, one would expect the answer is affirmative.

We noticed that the correlation between SiMNP and DIRECT-zol was lower than that between SiMNP and TRIzol (compare **Fig. 3A and 3C**). In the case of SiMNP vs. TRIzol, each sample was divided equally between SiMNP and TRIzol protocols and processed in parallel. On the other hand, the samples used in the SiMNP and DIRECT-zol comparison was extracted at different times. This was due to clinical reasons. Priority was given to extract enough RNA for NGS using the DIRECT-zol kits. As a results, samples were not equally divided between the SiMNP and DIRECT-zol extractions, and the swab might be present in one but not the other extraction method. These reasons could contributed to the lower correlation between the two methods. Nonetheless, evidence from 36 clinical samples, 2 wastewater samples and 1 pathogens control sample showed that MAVRICS ravels performance of commercial reagents.

We noticed an interesting lack of correlation between the amount of total RNA and viral RNA (**Supplementary Fig. 2A-C, 2D-G, Supplementary Fig. 3D vs. Fig. 3A-B**). For example, RNA concentration of S667 was below the detection range of Qubit fluorometer, and yet the copy number of SARS-CoV-2 was higher than S659, which had the one of the highest RNA concentration (**Supplementary Fig. 3B-C**). SiMNP tends to have lower total RNA yield, but has lower Ct values when compared to other methods (**Supplementary Fig. 3C**). There could be at least two possibilities. First, SiMNPs may favor binding of RNA similar to the viral RNA. This could be due to the surface chemistry or high surface area to mass ratio of nanoparticles. Second, SiMNPs may be more efficient in removing contaminants that inhibit reverse transcription and PCR. The exact reasons for this phenomenon need to be further studied.

In summary, we developed MAVRICS to enable safe, economical and effective extraction of RNA from clinical and environmental samples. The performance of MAVRICS rivals commercial RNA extraction kits in validated diagnostic tests of SARS-CoV-2, influenza viruses and respiratory syncytial virus. MAVRICS has the potential to become an enabling technology for widespread community testing and wastewater monitoring in the current and future pandemics.

## Data Availability

All data referred to in the manuscript are available upon request.

https://www.protocols.io/view/marvics-a-robust-and-safe-magnetic-nanoparticle-ba-bik4kcyw?step=6

## Acknowledgements

We thank KAUST Rapid Research Response Team (R3T) for supporting our research during the COVID-19 crisis. We thank members of the KAUST R3T for generously sharing materials and advices. We thank Professor Imed Gallouzi of McGill University for the useful discussion. We thank members of the Li laboratory, Chongwei BI, Baolei Yuan, Xuan Zhou, Samhan Alsolami, Yingzi Zhang, and Yeteng Tian, for helpful discussions; Marie Krenz Y. Sicat for administrative support. We thank members of the Pain lab for technical assistance.

## Author contributions

ML and GRM performed majority of the molecular biology experiments. ML and GRM analyzed the data and wrote the manuscript. RS, SM, and JX performed experiments. AR and PH performed experiments on wastewater. FSA, AK, AMH, NAMA and AA collected clinical samples. SH and AP coordinated the clinical samples and molecular testing. ML conceived and supervised the study.

## Supplementary Materials

**Supplementary figures 1 to 3**

**Supplementary protocol 1**

**Supplementary Fig. 1.**
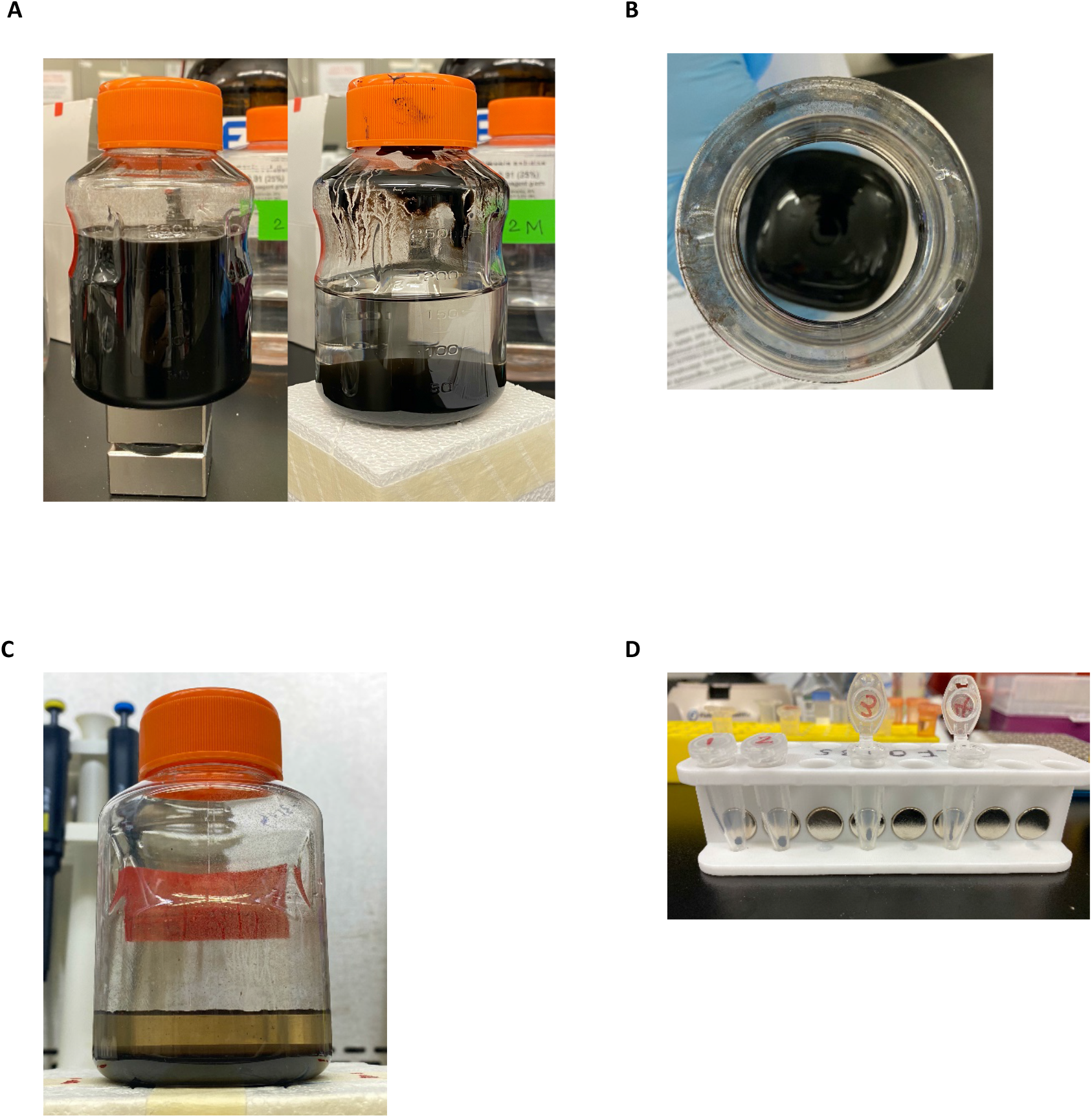
SiMNP synthesis. **A**. Pictures of the synthesized MNPs before (left) and after (right) pelleting by a strong magnet. **B**. A picture of the MNP pellet after decanting the solution. **C**. SiMNPs stored in equal volume of RNase-free water. **D**. A picture of a simple magnetic tube rack used to pellet the SiMNPs in Eppendorf tubes.

**Supplementary Fig. 2.**
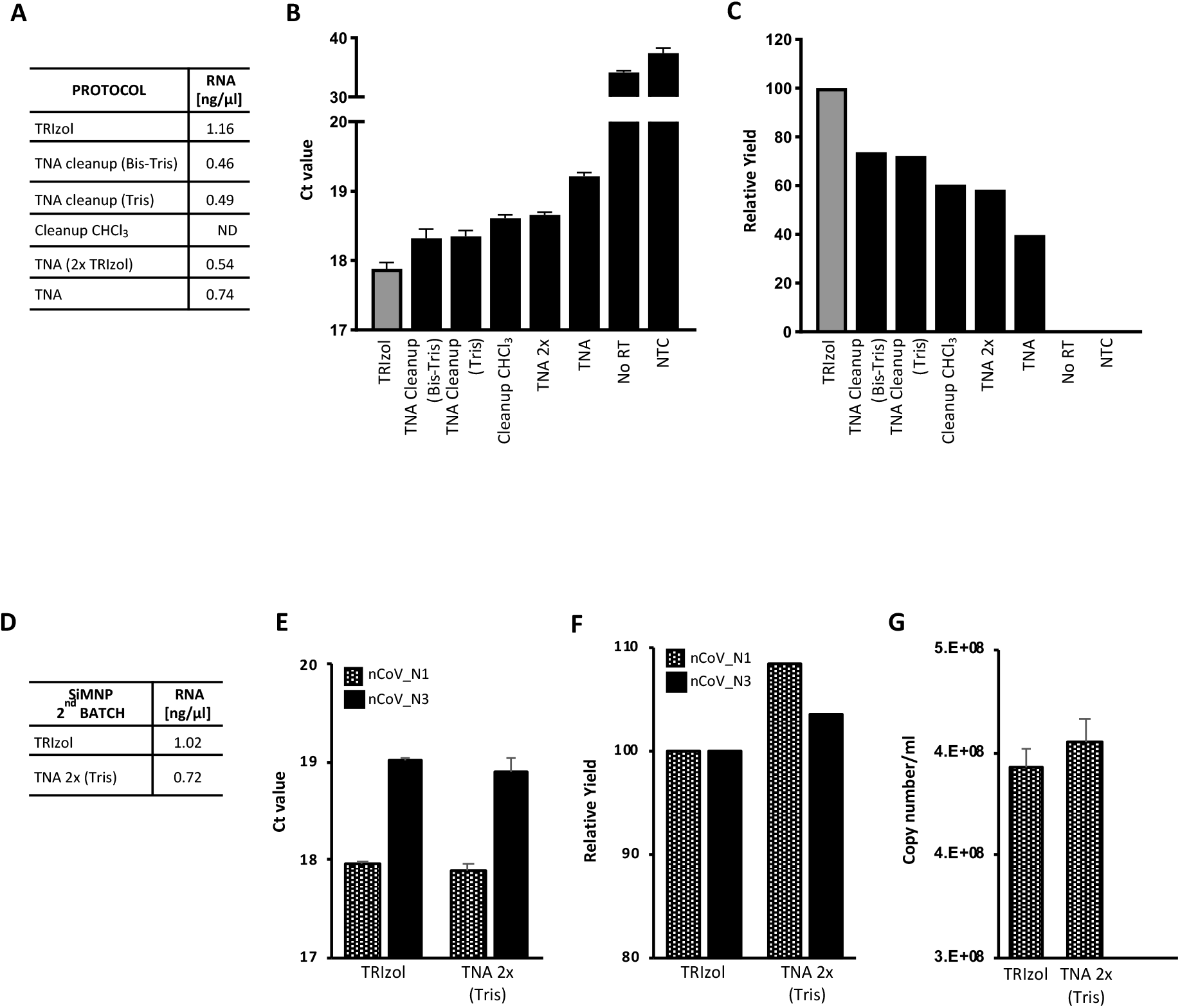
Optimization of SiMNP protocol to improve viral RNA extraction. After extraction using TRIzol or different SiMNP protocols RNA was quantified (**A**). **B-C**: the SARS-CoV-2 RNA recovery was compared using the 2019-nCoV_N1 probe rRT-PCR assays (**B:** Ct values, **C**: viral RNA yield relative to TRIzol extraction). **D-F**: Similar to **A-C** but using a second synthesis of SiMNPs. In the experiment using the second batch of SiMNP, the copy number of SARS-Cov-2 RNA in the original sample calculated by the standard curve method (**G**). Tris: Tris-HCl pH6.5 buffer. Bis-Tris: Bis-Tris, pH6.5 buffer, TNA 2x: TNA protocol with an additional TRIzol wash. N=3, error bars represent standard errors.

**Supplementary Fig. 3.**
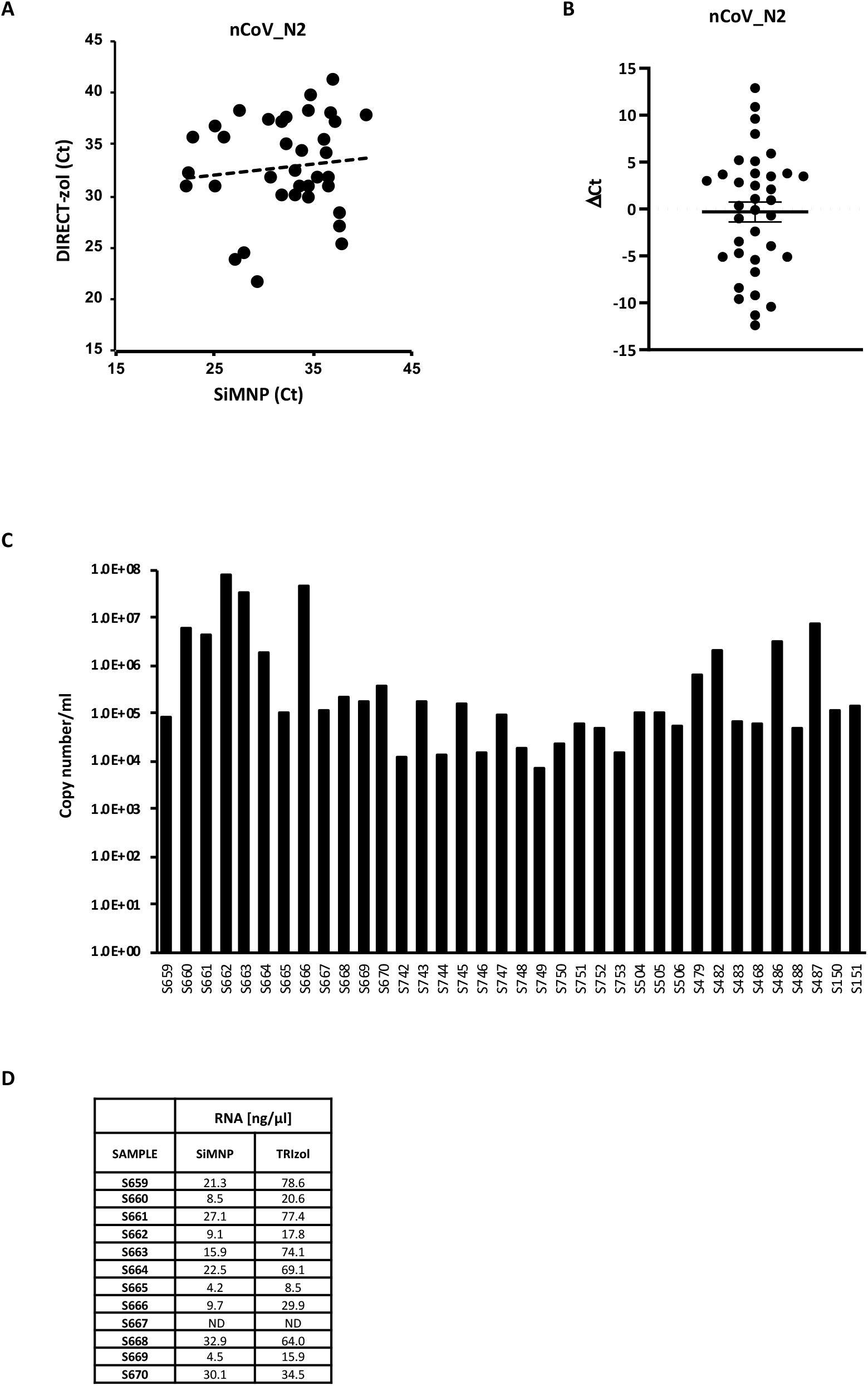
RNA extraction from CIVID-19 clinical samples and comparison between MAVRICS and commercial kits. **A-B**. RNA extraction was done from 36 samples using MAVRICS or DIRECT-zol protocol and the SARS-CoV-2 RNA recovery was compared using the 2019-nCoV_N2 probe. The graphs show the correlation between Ct values (**A**) and ΔCt value (**B**, mean and standard errors are shown). **C**. SARS-CoV-2 viral load (Copy number/ml) of 36 samples obtained using the MAVRICS protocol. **D**. Total RNA yield by SiMNP and TRIzol protocol. RNA concentration was evaluated by the Qubit fluorometer method. N=3.

### Supplementary protocol 1. Magnetic nanoparticle synthesis, silica coating and RNA extraction

Silica magnetic nanoparticles (SiMNP) synthesis

1. Core magnetic nanoparticle synthesis and silica coating of MNPs were done following published protocols (Protocols 1.1 and 2.1 in reference 12).

Preparation of Bis-Tris Buffer (50 mL)

1. Dissolve 14.33 g guanidinium hydrochloride and 104.6 mg Bis-Tris in 45 mL of 100% ethanol. NOTE: Add 40 ml of 100% ethanol to the other chemicals, and wait for guanidinium hydrochloride to completely dissolve and add the remaining volume of 100% Ethanol.
2. Adjust pH (<6.5) with HCl, and adjust the volume with H_2_O to 50 mL.

Sample preparation

1. Oropharyngeal or nasopharyngeal swabs were steeped in 1 mL acid guanidinium thiocyanate-phenol-chloroform (AGPC, e.g., TRIzol Reagent or TRI reagent).

RNA extraction

1. In an Eppendorf tube, add 200 µl clinical sample and 200 µl Bis-Tris buffer, mix well by vortexing.
2. Add 40 µl SiMNP, mix 5 min at 1300 rpm.
3. Spin the tube for 2-3 seconds, settle the SiMNPs on a magnetic stand and remove the supernatant. Remove the tube from the magnetic stand.
4. Mix 200 µl of AGPC (TRIzol or TRI reagent) and 200 µl Bis-Tris buffer, add to the SiMNPs, mix well by vortexing.
5. Settle the SiMNPs on a magnetic stand and remove the supernatant. Remove the tube from the magnetic stand.
6. Add 400 µl of 90% ethanol, spin for 2-3 seconds, settle the SiMNPs on a magnetic stand and remove the supernatant. Remove the tube from the magnetic stand.
7. Repeat Step 6 three more times for a total of 4 ethanol washes.
8. After removing the supernatant from the last ethanol wash, dry the SiMNPs on a heat block at 50°C. Keep the lid open, no shaking. Do not elute before the SiMNPs are dried.
9. To elute the RNA, add 40 µl nuclease-free water, and mix 5 min at 1300 rpm at room temperature.
10. Settle the SiMNPs on a magnetic stand and transfer the eluted RNA to a new RNase-free tube.
11. Analyze RNA concentration and purity using a Qubit fluorometer or Nanodrop.
12. Store at −80°C or use immediately.

Reverse transcription (RT) and Real-time PCR

1. RT: use 4 µl of eluted RNA and follow the instructions for SuperScript™ IV Reverse Transcriptase adding the RNase H incubation step.
2. Real-time PCR: For each 10 µl qPCR reaction mix 1.5 µl cDNA, 0.5 µl SARS-CoV-2 (2019-nCoV) CDC qPCR Probe Assay, 5 µl TaqMan Fast Advanced Master Mix, and 1.5 µl nuclease-free water. Run qPCR on a Biorad CFX384 Touch Real-Time PCR Detection System (or similar instrument) using the following program: 50°C for 2 min, 95°C for 2 min followed by 45 cycles of 95°C for 5 sec and 59°C for 30 sec.

